# The impact of Indigenous American-like ancestry on risk of acute lymphoblastic leukemia in Hispanic/Latino children

**DOI:** 10.1101/2025.01.14.25320563

**Authors:** Jalen Langie, Tsz Fung Chan, Wenjian Yang, Alice Y. Kang, Libby Morimoto, Daniel O. Stram, Nicholas Mancuso, Xiaomei Ma, Catherine Metayer, Philip J. Lupo, Karen R. Rabin, Michael E. Scheurer, Joseph L. Wiemels, Jun J. Yang, Adam J. de Smith, Charleston W.K. Chiang

## Abstract

Acute lymphoblastic leukemia (ALL) is the most common childhood cancer, with Hispanic/Latino children having a higher incidence of ALL than other racial/ethnic groups. Among genetic variants previously implicated in ALL risk, a number of them were found to be enriched in Indigenous American (IA)-like ancestries and inherited by many Hispanic/Latino individuals. However, due to potential confounding from environmental factors, the association between IA-like ancestry and risk for ALL has remained unclear. In this study, we characterized the impact of IA-like ancestry on overall ALL risk and on the frequency and effect size of known risk alleles, while accounting for non-genetic correlates of ancestry. Contrary to previous findings, we found that global IA-like ancestry was not significantly associated with ALL risk after adjusting for socio-economic indicators. On the other hand, locally at known ALL risk regions, we uncovered that increasing copies of the IA-like haplotype were positively and significantly associated with ALL risk (*e.g.* the IA-like haplotype had ∼1.33 times the odds of harboring the risk allele compared to non-IA-like haplotypes), but we found no evidence of interaction between genotype and ancestry in relation to ALL. Admixture mapping identified replicable association signals at chr7p12.2 and chr10q21.2, consistent with the benefit of leveraging genetic ancestry in identifying genetic risk loci. Taken together, our results suggest that increased risk of ALL in Hispanic/Latino children may be conferred by higher frequency of risk alleles within IA-like ancestry, and that local ancestry-based analyses are robust stratagems to elucidate genetic etiology of disease.

## Introduction

Acute lymphoblastic leukemia (ALL) is the most common childhood cancer and accounts for 25% of malignancies before the age of 20 (ref. ^1^). The long-term survival rate of the disease in children is upwards of 85-90% ^2^, but survival rate decreases to approximately 40% after relapse^3–6^ and 30-40% in individuals over the age of 35. Despite decades of investigation into ALL risk, the etiology of the disease is not well understood and disparities in risk persist. Children of Hispanic/Latino ethnicity have 1.4 and 2 times the risk of developing ALL compared to their Non-Latino White and Black counterparts, respectively^7^. This disparity in risk widens in margin for Hispanic/Latino adults^7^ and is specific to the B-cell immunophenotype – which comprises approximately 85% of cases^8^. While environmental factors are involved in this increased risk, genetic factors, which have been historically understudied in this population^9,10^, likely contribute in part to this disparity^11,12^

Genomic, migrational, and ecological data strongly suggest a complex genetic etiology of ALL, both broadly across humans and within diverse Latin American populations. Most directly, previous genome-wide association studies (GWAS) have estimated the heritability of ALL to be approximately 18-21% in both Hispanic/Latino and Non-Latino White populations, and approximately 24% of familial relative risk has been attributed to known ALL risk variants^13–15^. Consistent with these estimates, GWAS have identified a number of loci associated with ALL susceptibility, including risk alleles located near genes such as *IKZF1*, *ARID5B*, *GATA3*, and *ERG* that appear to have higher frequency or stronger effects in Hispanic/Latino populations^12,16–18^. Previous migration studies have indicated that non-native- and US-born Hispanic/Latino individuals have comparable rates of B-cell ALL to each other, and both have higher (∼2.13 times) incidence rates compared to their Non-Latino White counterparts^19,20^. Furthermore, ecological data has indicated that countries with the highest age standardized incidence rates of ALL were in Latin America (*i.e.* Honduras and Mexico), while countries with the lowest incidence rates were in Africa, with European countries exhibiting intermediate incidence^21,22^.

Notable amongst populations with differing incidence rates of ALL is the presence or dominance of a particular ancestry component. Due to the unique demographic history of Latin America, Hispanics/Latino populations are admixed - drawing ancestral components from the American, European, and African continents^23,24^. Previous literature has alluded to associations between Indigenous American (IA)-like ancestry and ALL, suggesting that every 20% increase in IA-like ancestry was associated with a 20% increase in odds of developing B-cell ALL^25^. Because Hispanics/Latinos inherited a significant proportion of their genetic ancestries from Indigenous Americans (though there is geographical heterogeneity across the Americas), this association was presumed to explain the migration and ecological findings above. However, global ancestry can be confounded by non-genetic factors such as socio-economic status^26^, which was not considered in previous analyses^25^. Further exploration of the association between IA-like ancestry and ALL risk would need to account for this potential confounding.

Given the important role of genetic factors and genetic ancestry in ALL risk, we undertook a systematic admixture analysis in the largest ALL cohort of self-reported Hispanic/Latino children to date. Here, we sought to clarify the association between IA-like ancestry and ALL risk through a study design that would better account for non-genetic confounders and leveraged local ancestry analyses, which are less susceptible to confounding by environmental factors and have previously yielded promising results in examining ancestry-ALL relationships^30^. We investigated both inferred global ancestry across the genome and local ancestry at known ALL loci for associations with risk. Further, we employed admixture mapping^27^, which is best powered to detect loci that exhibit frequency differences between ancestries and may underlie the risk differences between populations^28,29^.

We note that, given our goal to further understand the disparity in ALL risk epidemiologically observed among Hispanic/Latino children, we inferred the genetic ancestries and characterized their associations with ALL risk in self-reported Hispanic/Latino individuals. We defined genetic ancestries at the continental (e.g. Africa, Europe, and Americas) level, which represents patterns of genetic similarities across geographical space over the last ∼10-50k years due to human migrations and isolation-by-distance. These relatively distinct components of ancestries, though arbitrarily defined, are only recently reunited in admixed populations, such as in individuals of Hispanic/Latino origin. These components are inferred and defined relative to reference populations available to us and are leveraged to uncover associations with traits in this research setting but are invariably a simplified representation of the complex nature of genetic ancestries. We thus append the suffix “-like” to refer to these ancestries to remind the reader of this simplification and uncertainty^30^.

## Methods

### Study Subjects and Quality Control

We use the term “Hispanics/Latinos” as a population descriptor of our study subjects to align with both the self-reported race/ethnicity categories and the definitions of the USA Office and Management Budget^31^, while acknowledging that we focus on individuals of Latin American origin. Our analysis utilized the following cohorts of self-reported Hispanic/Latino study participants: the California Cancer Linkage Project (CCRLP), the Kaiser Resource for Genetic Epidemiology Research on Aging Cohort (Kaiser-GERA), the California Childhood Leukemia Study (CCLS), Children’s Oncology Group (COG), Multi-Ethnic Study of Atherosclerosis (MESA), the Adolescent and Childhood Cancer Epidemiology and Susceptibility Service for Texas and Reducing Ethnic Disparities in Acute Leukemia (ACCESS/REDIAL), and the Hispanic Community Health Study/Study of Latinos (HCHS/SOL). All relevant information on sample size, arrays, study design, and references/dbGaP accession numbers can be found in **Table 1**. Following the practice in previous studies^12,13,32^, cases from CCRLP were augmented with external controls from Kaiser-GERA genotyped on a similar array. Cases from COG were paired with external controls from MESA and ACCESS/REDIAL cases were paired with external controls from HCHS/SOL. HCHS/SOL was a multi-center study, with study participants being ascertained from Bronx, New York; Chicago, Illinois; Miami, Florida; and San Diego, California. The SOL San Diego center was chosen as controls for ACCESS/REDIAL due to similarity in the first 2 principal components and estimated ancestry distributions (**Supplementary** Figures 1-2).

**Table 1:** All cohorts included in the analysis. Information includes title, design, sample size, genotype array, dbGaP accession number or reference, and how each study was used in our analysis.

| Study | In-Text Name/<br>Acronym | Study Design | N | Genotype Array | Reference or<br>Accession Number | Usage |
| --- | --- | --- | --- | --- | --- | --- |
| **California Cancer Linkage Project** | CCRLP | Population Based case-control study | 4033 | Affymetrix Axiom World Array | Wiemels et al., 2018 (ref <sup>32</sup>) | Global and local ancestry analyses; Discovery Admixture Mapping Cohort (combined with Kaiser-GERA) |
| **Kaiser Resource for Genetic Epidemiology Research on Aging Cohort** | Kaiser-GERA | Cohort study (controls only) | 6417 | Affymetrix Axiom World Array | phs000788.v1.p2 | Discovery Admixture Mapping Cohort (combined with CCRLP) |
| **California Childhood Leukemia Study** | CCLS | Population Based case-control study (cases and controls) | 1120 | Illumina OmniExpress v.1 | Ma et al., 2004 (ref <sup>33</sup>) | Global and local ancestry analysis; Replication cohort |
| **Children's Oncology Group (AALL0232, COG9904/9905)** | COG | Clinical Trial (cases only) | 836 | Genome-Wide Human SNP Array 6.0 | phs001350.v1.p1 | Replication Cohort (combined with MESA) |
| **Multi-Ethnic Study of Atherosclerosis** | MESA | Cohort Study (controls only) | 1118 | Genome-Wide Human SNP Array 6.0 | phs000209.v13.p3 | Replication Cohort (combined with COG) |
| **Adolescent and Childhood Cancer Epidemiology and Susceptibility Service for Texas and Reducing Ethnic Disparities in Acute Leukemia** | ACCESS/<br>REDIAL | Hospital Cases (cases only) | 510 | Illumina Global Screening Array | Harris et al., 2023 (ref <sup>34</sup>) | Replication Cohort (combined with HCHS/SOL) |
| **Hispanic Community Health Study/Study of Latinos** | HCHS/SOL | Cohort Study (controls only) | 2285 | Illumina Omni2.5M array | phs000810.v2.p2.c2;<br>phs000810.v2.p2.c1 | Replication cohort (combined with cases from Texas) |

All analyses were limited to autosomes. Within each study, SNPs and individuals were removed based on the following pre-defined quality control thresholds: SNP call rate < 0.98, MAF < 0.01, *p_HWE_* < 1 × 10^-5^, high relatedness 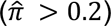, Genome Heterozygosity Rate > 6 standard deviations from the mean, or sample call rate < 0.95. Palindromic SNPs were also excluded as they could impact imputation accuracy due to unidentified strand flip or inversion between genome build^35^. The number of individuals and SNPs remaining can be found in **Supplementary Table 1**. Local ancestry was inferred using RFMIX^36^. Reference haplotypes were obtained from the genome aggregation database (gnomAD)^37^ version 3, including 708 African, 671 Non-Finnish European, and 94 Indigenous American individuals previously chosen for ancestry inference in Hispanics/Latinos^38^. After restricting to the intersection of SNPs across each paired dataset, each dataset was imputed separately using the TopMed Imputation Server^39^. We observed systemic differences in imputation quality as measured by the imputation R^2^ between cases and external controls, presumably due to batch effects in data generation or subtle differences in ancestry compositions between cohorts and differential imputation accuracies across populations^40^. Consequently, for all comparisons of cases to controls, we imposed filters such that only SNPs with an imputation R^2^ > 0.30 in both cases and controls and a R^2^ difference < 0.15 between cases and controls were included in the analysis.

### Global Ancestry Analyses

Global ancestry for each study participant was calculated by averaging local ancestry estimates across the genome. We performed logistic regression of ALL case-control status on global IA-like ancestry in CCRLP and CCLS, studies in which indicators of socio-economic status (SES) variables were available. Covariates included African-like ancestry, sex, and SES indicators (highest education obtained by a parent and annual household income). Income was categorized in 6 strata, in accordance with its treatment in CCLS^41^ : <$15,000, $15,000-$29,999, $30,000-$44,999, $45,000-$59,999, $60,000-$74,999, and >$75,000. In CCLS, highest educational attainment of either parent was divided into 4 categories: Less than elementary school, high school, some college, and a bachelor’s degree or higher. In CCRLP, the strata were less than 9^th^ grade, 9^th^-12^th^ grade, and some college or more. Both SES indicators were treated as ordinal categorical variables, in which the lowest category was used as a reference. Linearity assumptions were assessed using residual and QQ plots. Analysis was carried out using R v.4.3.1 (ref. ^42^).

### Enrichment of risk allele on IA-like ancestry haplotypes

We performed local ancestry analyses at known ALL loci, based on the list provided in ref. ^11^. We excluded the T-cell ALL-specific association locus at *USP7*^43^ given that the increased risk of ALL in Hispanics/Latinos is limited to B-cell ALL. We restricted analyses to remaining SNPs that were independent at the local ancestry level (*i.e.* choosing SNPs that were not in admixture linkage disequilibrium; N=20) using PLINK’s clumping algorithm^44^. SNPs were clumped if the diploid local IA-like ancestry assignments covering the SNPs were 90% correlated and within 10 Mb of each other.

A total of three analyses were performed using local ancestry estimates. First, across known ALL loci^11^, we cross-tabulated local genetic ancestry (number of copies of the IA-like, African-like, or European-like haplotypes) and risk allele dosage to identify association of risk alleles with ancestry. To establish a null distribution to which we would compare our observed ancestry-risk allele association, we randomly selected 1000 sets of 20 frequency-matched alleles from across the genome, without replacement. Differences between the null and known loci distributions were evaluated via a Chi-Squared Goodness of Fit test. We restricted this analysis to CCRLP and CCLS controls, to approximate the frequency of the risk allele in the general population. Second, we performed a logistic regression of ALL case-control status on IA-like ancestry dosage at each locus, including as covariates global African-like and European-like ancestry, and sex, to evaluate how likely is the IA-like ancestry associated with increasing ALL risk. Third, to test for presence of heterogeneous effect sizes across ancestries, we performed logistic regression of ALL case-control status on risk allele genotype, IA-like ancestry dosage, sex, the first 20 principal components, and an interaction term of local ancestry and risk allele dosage. Analyses were carried out either using PLINK v2.0 (ref. ^45^) or R v.4.3.1 (ref. ^42^).

### Admixture mapping, regional association testing, and replication

We performed a genome-wide scan for loci in which IA-like ancestry was associated with ALL risk. We regressed ALL case-control status on the local IA ancestry dosage, global African-like and European-like ancestry, sex, and the first 20 principal components. We performed this analysis in CCRLP/Kaiser as our discovery dataset. The significance threshold for genome-wide admixture mapping was 5.7 x 10^-5^, as suggested in literature previously for Hispanic/Latino populations^46,47^.

We fine-mapped putative admixture association region through single-variant association testing in the same dataset. The admixture peak region^48^ was defined as a range of positions that contains all SNPs with a -log10(*P*) between -log10(*P*_*am*_) and -log10(*P*_*am*_) – 1, with *P*_*am*_ representing the p-value associated with the lead SNP from admixture mapping. Genotypes of all imputed SNPs within this region passing QC filtering were tested via logistic regression, with sex and the first 20 principal components as covariates. Significance threshold for each region were computed using 1000 rounds of permutation of case-control labels, choosing the threshold that controls for the 5% false discovery rate. We attempted to replicate the association at the lead variants from regional association testing and conditional analysis in additional Hispanic/Latino cohorts: CCLS, COG/MESA, and SOL-SD/REDIAL. The replication cohorts were meta-analyzed together by inverse variance weighting in the software *METAL*^49^.

### Evaluating the impact of using self-reported race/ethnicity labels versus genetic inferred ancestry

We opted to perform admixture mapping using all self-reported Hispanic/Latino individuals to be inclusive and avoid forming study exclusion criteria based on estimated genetic ancestry. However, alternative approaches could choose to include participants based on inferred genetic ancestries. To examine, in our context, the impact of informing study inclusion based on genetically inferred ancestry, we implemented a software, Harmonizing Ancestry and Race Ethnicity^50^ (HARE; Fang et. al, 2019) on genetic data from CCRLP/Kaiser-GERA (**Supplementary** Figure 3. HARE is a two-step machine learning-based algorithm that identifies individuals who may have discordant self-identified race/ethnicity (SIRE) and genetically inferred ancestry (GIA) information, given the mapping of the two learned from the rest of the dataset. The first step is a training phase in which genetically inferred ancestry based on principal components are used to predict available self-reported race/ethnicity in a support vector machine (SVM). The second step is HARE strata assignment, in which the highest probability of being assigned to a stratum is contrasted with the probability that the individual belongs to their self-reported stratum using preset thresholds. If the ratio of the probabilities exceeds a pre-defined threshold, an individual with existing SIRE could be assigned as missing (*i.e.* removed from analysis), while an individual without SIRE would be assigned into the top predicted strata.

We applied HARE with the default parameters and thresholds^50^, training a multi-class SVM with a radial kernel on the first 30 principal components (calculated using PLINK^45^ v2.0) of genotype data from individuals across all self-reported race/ethnicity groups from CCRLP/Kaiser-GERA (10,450 Hispanic/Latino, 2,191 Non-Latino African, 5,335 Non-Latino East Asian, 58,503 Non-Latino European individuals). We first evaluated if the HARE algorithm would exclude any study participants due to discordance between SIRE and GIA. We also assessed the rate of discordance between SIRE and GIA by performing 10-fold cross validation in which we randomly masked the SIRE information of 10% of study participants for each race/ethnicity group in each trial. This assessment simulates a scenario where one may define study exclusion criteria based on comparing the GIA to a known reference panel. Percent discordance was defined as the number of individuals classified as missing or as a stratum not corresponding to the masked SIRE label. We then investigated the impact of this study exclusion criteria on our admixture mapping result.

### IRB Statement

This study was approved by the institutional review boards of the California Health and Human Services Agency, the University of Southern California, Children’s Oncology Group, Baylor College of Medicine, Yale University, and University of California, Berkeley. All study participants were de-identified and newborn bloodspots from the CCRLP were obtained via a consent waiver from Committee for the Protection of Human Subjects of the State of California.

## Results

### The association between global Indigenous American-like ancestries, ALL risk, and SES indicators

Previous studies have identified a significant and positive association between global IA-like ancestries and ALL risk^25^ (Walsh et al., 2013) but did not adjust for socioeconomic status (SES) variables that often confound ancestry-disease associations. To quantify the effects of IA-like ancestry on ALL risk, we tested the association between ALL case-control status and global IA-like ancestry, while adjusting for African-like ancestry, sex, as well as covariates that are proxies for SES, such as highest parental education and annual household income. We performed these assessments for all self-reported LAT individuals in both CCRLP (excluding Kaiser-GERA controls; N = 4033) and CCLS (N = 605), as both of these cohorts have available SES variables, contained internal controls and are less likely to be confounded due to their study design. In CCLS, IA-like ancestry was significantly associated with ALL risk, prior to including SES indicators in the model, as previously reported (Walsh et al., 2013). In the base model adjusting for sex and African-like ancestry, a 20% increase in IA-like ancestry was associated with a 23% (OR: 1.23, 95% CI: 1.07-1.42; *P* = 0.003) increase in odds of developing ALL. However, this association appears confounded, as inclusion of SES covariates strongly attenuated the strength of association (OR: 1.12; 95% CI: 0.96-1.31) and case/control status is no longer significantly associated with IA-like ancestry (*P = 0.*159; **Supplementary Table 2**). In CCRLP, IA-like ancestry was not significantly associated with ALL risk in the base model adjusting for only African-like ancestry and sex, and did not qualitatively change when SES covariates were included (**Supplementary Table 2**). Regression analysis between the SES variables and ALL case-control status and ancestry, respectively, further supported the suggestion that socio-cultural factors can act as confounders in ancestry-trait relations. For instance, we observed higher income to be associated with lower risk of disease (*e.g.* 0.27 – 0.76 times the odds of developing ALL; **Supplementary Tables 3-4**), and higher European-like ancestry is associated with higher socio-economic status (**Supplementary Tables 5-6**). Together, because SES measures can correlate with both ancestry and disease risk, the previously observed association between IA-like ancestry and ALL risk (Walsh et al., 2013) may have been confounded.

### Known ALL risk loci are enriched among Indigenous American-like ancestral haplotypes

While global ancestry association analyses can be confounded by socioeconomic indicators, use of local ancestry to identify associations at risk loci may be more robust. Further, the absence of an association between IA-like ancestry and ALL risk globally would not necessarily preclude an association locally at known risk loci. Thus, we performed three analyses leveraging the pattern of local ancestry at known B-cell ALL risk loci^11^ (**Supplementary Table 7**) to examine if the risk alleles are found more frequently or show larger effects on IA-like ancestry haplotypes. First, we tabulated the proportion of times the risk allele at known ALL loci is associated with the African-like, IA-like, or European-like ancestry haplotypes, and compared these to the proportions found across 1000 sets of SNPs randomly selected across the genome and matched by the risk allele frequencies. Across a total of 20 quasi-independent ALL SNPs (**Methods**), the risk alleles were found on the African-like, IA-like, and European-like ancestry haplotypes 5.76%, 45.35%, and 48.88% of the time, respectively, in CCRLP. This significantly deviates from the genome-wide expectation (4.83%, 43.33%, and 51.85% for African-like, IA-like, and European-like ancestry haplotypes; p-value = 1.04x10^-36^ by a chi-square goodness-of-fit test), with IA-like haplotypes showing a significant enrichment in association with the risk alleles. We also replicated this observation in CCLS, with IA-like haplotypes enriched with ALL risk alleles (p-value = 1.629x10^-9^). Consistent with this finding, the IA-like haplotypes have 1.32-1.33 (95% CI: 1.29 - 1.36 in CCRLP, 1.26-1.41 in CCLS) times the odds of carrying the risk allele compared to Non-IA-like haplotypes (p = 5.61x10^-88^ and 2.61x10^-23^ in CCRLP and CCLS, respectively; **Supplementary Table 8**).

In the second local ancestry analysis, we regressed ALL case-control status on the number of copies of IA-like ancestry segments locally at each known ALL locus, while adjusting for sex and global African and European ancestry proportions. Because we anticipate that risk alleles for ALL are found disproportionately on IA-like haplotypes across these loci, we would expect that IA-like ancestries show an enrichment of association increasing the risk of ALL in the setting of admixture mapping studies. Indeed, we found a positive association of IA-like ancestry with ALL at 14 out of 20 known B-cell ALL loci. At all loci in which IA-like ancestry was nominally significantly (p < 0.05) associated with ALL, IA ancestry was positively associated with risk (Binomial test, p = 0.008 **Table 2**). To confirm that the known risk alleles were in fact driving the ancestry association, we included risk genotype as an additional covariate in each model. At each locus, upon adjustment for risk genotype, the association between IA-ancestry and ALL risk reduced in significance (**Supplementary Table 9**). Taken together, our results suggest that IA-like haplotypes are enriched for association with increased ALL risk at known loci, driven by increased frequency of risk allele.

**Table 2:** Indigenous American-like ancestry is positively associated with risk at known ALL loci. ALL case-control status was regressed on local Indigenous ancestry dosage, African-like and European-like global ancestries, and sex^a^. Loci that reach nominal significance (P < 0.05) are in **bold**)

| Gene | Chr | Location | OR | L95 | U95 | P-value<sup>a</sup> |
| --- | --- | --- | --- | --- | --- | --- |
| BCL11A | 2 | 2p16.1 | 0.94 | 0.86 | 1.02 | 0.138 |
| RPL6P5 | 2 | 2q22.3 | 1.01 | 0.93 | 1.10 | 0.751 |
| C5orf56 | 5 | 5q31.1 | 1.07 | 0.98 | 1.17 | 0.113 |
| BAK1 | 6 | 6p21.31 | 0.946 | 0.87 | 1.03 | 0.208 |
| MYB/HBS1L | 6 | 6q23 | 0.99 | 0.91 | 1.07 | 0.749 |
| **IKZF1** | **7** | **7p12.2** | **1.32** | **1.21** | **1.43** | **2.990 x 10<sup>-10</sup>** |
| CCDC26 | 8 | 8q24.21 | 0.93 | 0.85 | 1.01 | 0.084 |
| CDKN2A/2b | 9 | 9p21.3 | 0.93 | 0.86 | 1.02 | 0.115 |
| TLE1 | 9 | 9q21.31 | 1.04 | 0.96 | 1.13 | 0.358 |
| **GATA3** | **10** | **10p14** | **1.14** | **1.04** | **1.24** | **0.003** |
| **BMI1/PIP4K2A** | **10** | **10p12.2/10p12.31** | **1.16** | **1.07** | **1.27** | **5.422 x 10<sup>-4</sup>** |
| **ARID5B** | **10** | **10q21.2** | **1.29** | **1.19** | **1.41** | **4.924 x 10<sup>-9</sup>** |
| **JMJD1C** | **10** | **10q21.3** | **1.27** | **1.16** | **1.38** | **5.383 x 10<sup>-8</sup>** |
| **TET1** | **10** | **10q21.3** | **1.15** | **1.06** | **1.25** | **0.001** |
| LHPP | 10 | 10q26.13 | 0.97 | 0.89 | 1.06 | 0.508 |
| **ELK3** | **12** | **12q23.1** | **1.10** | **1.01** | **1.19** | **0.038** |
| **CEBPE** | **14** | **14q11.2** | **1.10** | **1.01** | **1.20** | **0.022** |
| IKZF3 | 17 | 17q21.1 | 1.03 | 0.95 | 1.11 | 0.515 |
| IGF2BP1 | 17 | 17q21.32 | 1.02 | 0.93 | 1.11 | 0.728 |
| ERG | 21 | 21q22.2 | 1.02 | 0.93 | 1.11 | 0.711 |

In addition to ALL risk alleles being found more prevalently on IA-like haplotypes, risk alleles may also exert larger (or smaller) effects on ALL when they are found on the IA-like background. Therefore, for the third local ancestry analysis, we tested for effect size heterogeneity by including a SNPxAncestry interaction term in the standard association testing model. After Bonferroni correction, no significant interactions between genotype and local IA-like ancestry were observed in either CCRLP or CCLS, or when the two cohorts were combined (**Supplementary Table 10**). We additionally found no evidence of interaction between global IA ancestry and genotype (**Supplementary Table 11**). Therefore, we found no evidence that ALL risk alleles may exert larger effects on an IA haplotype than on other haplotypic backgrounds.

*Admixture mapping proposes putative novel ALL loci associated with Indigenous American-like ancestry*.

Although results from our global ancestry analysis showed minimal association between IA-like ancestry and ALL risk after adjusting for SES indicators, it does not preclude the possibility that some risk alleles show drastic differences in frequency between ancestries, and thus are amenable to being discovered through admixture mapping, particularly since we observed that IA-like ancestries are enriched for association with risk alleles for ALL (**Table 2** above). We thus performed admixture mapping using the CCRLP and Kaiser/GERA Hispanic/Latino cohort as our discovery panel (1930 cases and 6417 controls). Our discovery admixture mapping analyses identified genome-wide significant (threshold = 5.7 x 10^-5^; see **Methods**) signals at 2q21.2, 7p12.2, 10q21.2, and 15q22.31. The loci on chromosomes 7 (*IKZF1*) and 10 (*ARID5B*) are known, while those on chromosomes 2 and 15 are potentially novel (**Figure 2**).

**Figure 2:**
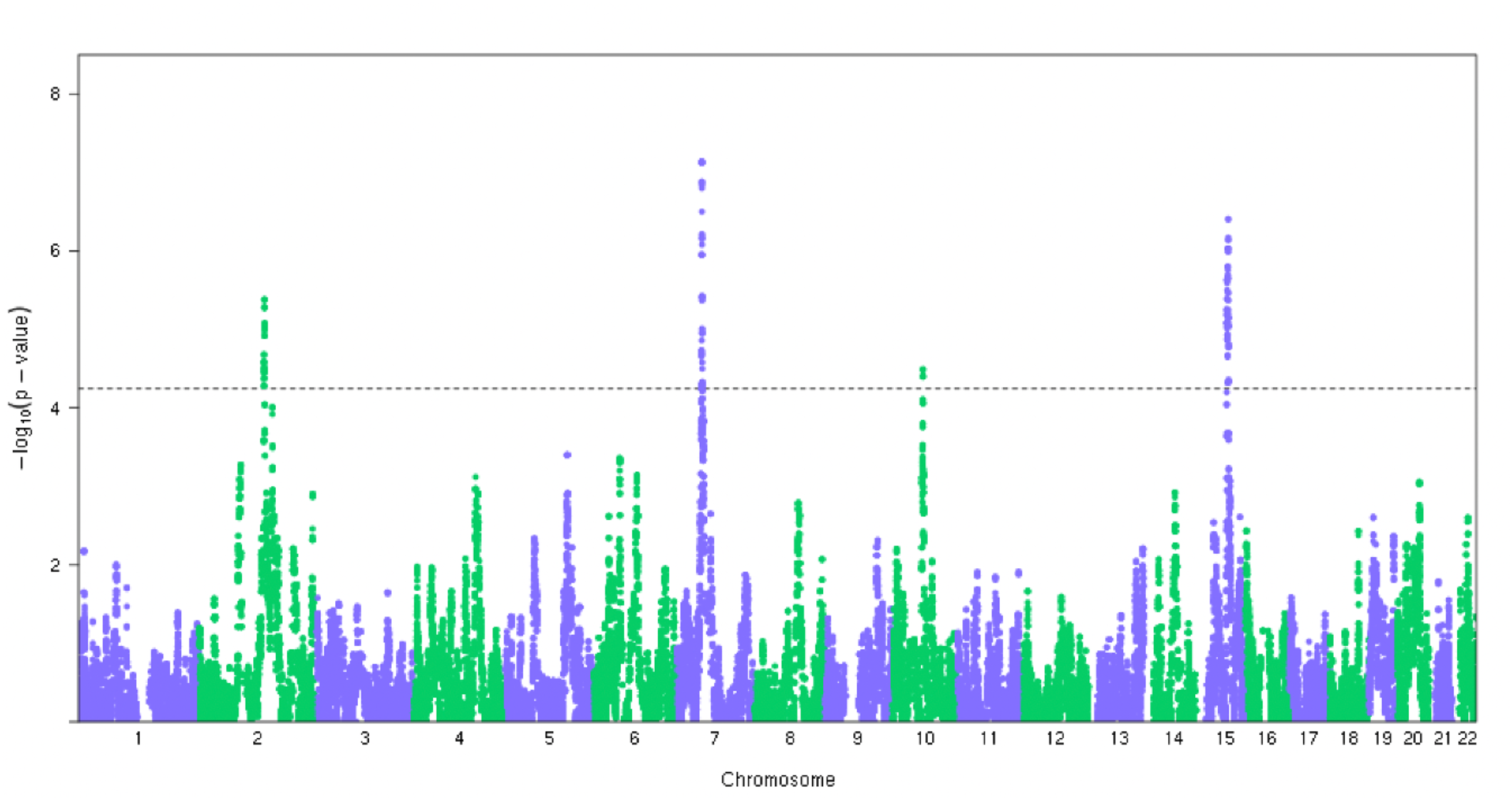
Manhattan plot of the admixture mapping analysis. The analysis identified signals at 2q21.2, 7p12.2, 10q21.2, and 15q22.31. The significance threshold is 5.7e-5.

We then fine-mapped each of the four admixture loci via single-variant regional association testing (**Methods**), which uncovered the putative lead SNPs in each region to be 2:133605881 (p = 4.209 x 10^-5^), 7:50395237 (p = 2.262 x 10^-12^), 10:61959980 (p = 2.245 x 10^-44^), and 15:66111790 (p = 3.011 x 10^-4^; **Table 3**). SNPs on chr2, 7, and 10 surpassed the regional significance threshold after accounting the number of tests performed in each region via permutation (**Methods**), and chr15 SNP was slightly below the threshold. These SNPs exhibit allele frequency differences by ancestry group (**Table 3**), and conditioning on these SNPs also generally attenuated the admixture mapping association (**Table 3)**. We sought replication of these associations in additional Hispanic/Latino cohorts, including cases from CCLS, COG, and REDIAL (see **Methods**), totaling 1,951 cases and 3,918 controls. In the meta-analyzed replication cohorts, only the lead SNPs or other SNPs in LD with the lead SNPs on chromosomes 7 and 10 were statistically significant (**Supplementary Table 12-13**). Through power calculations^51^, we found that, given observed odds ratios in the discovery cohort and combined allele frequencies in the replication datasets, our replication analysis had only 1.2% and 0.5% power to detect the associations on chromosomes 2 and 15, respectively; in contrast, we had 17.8% power to identify the signal on chromosome 7 and 100% power to detect the locus on chromosome 10.

**Table 3:** Odds Ratios, 95% CI’s, and p-values of top SNPs from regional association testing, under the admixture peak, as well as the crude and adjusted p-values of the admixture mapping tag SNP. The regional significance thresholds, determined via permutation to account for the regional multiple testing burden, are 7.906x10^-5^, 6.256x10^-4^, 1.905x10^-3^, and 1.270x10^-4^ for 2q21.2, 7p12.2, 10q21.2, and 15q22.31, respectively.*Risk allele frequencies (RAF) per ancestry group were obtained from European (Orcadian, Russian, Tuscan, French, Basque, Bergamo, Sardinian, Adygei), African (Bantu, Mbuti, Biaka, San, Yoruba, Mandenka), and Indigenous American (Surui, Karitiana, Maya, Pima, Colombian) superpopulation data from the Human Genome Diversity Project (accessed from the Genome Aggregation Database^38^ v4).

| Chr | Pos (Hg38) | rsID | Locus | Ref | Risk | $OR_{Top\ SNP}$<br>(95% CI) | $P_{Top\ SNP}$ | $RAF_{EUR}^*$ | $RAF_{AFR}$ | $RAF_{IA}$ | Gene | $P_{crude}$ | $P_{adj}$ | Peak<br>Region |
| --- | --- | --- | --- | --- | --- | --- | --- | --- | --- | --- | --- | --- | --- | --- |
| 2 | 134116620 | rs117345013 | 2q21.2 | T | C | 1.22 (1.11, 1.72) | $4.209 \times 10^{-5}$ | 0.010 | 0.000 | 0.390 | Unknown | $4.156 \times 10^{-6}$ | 0.005 | 133605881,<br>134236147 |
| 7 | 50390453 | rs6969942 | 7p12.2 | C | T | 1.35 (1.25, 1.47) | $2.262 \times 10^{-12}$ | 0.507 | 0.773 | 0.870 | IKZFI | $7.315 \times 10^{-8}$ | 0.002 | 50352094,<br>50457722 |
| 10 | 61959980 | rs4948492 | 10q21.2 | T | C | 1.85 (1.59, 2.13) | $2.245 \times 10^{-44}$ | 0.295 | 0.162 | 0.441 | ARID5B | $3.210 \times 10^{-5}$ | 0.202 | 61928110,<br>62306779 |
| 15 | 65877726 | rs12438510 | 15q22.31 | G | A | 1.23 (1.10,1.39) | $3.011 \times 10^{-4}$ | 0.000 | 0.000 | 0.340 | RAB11A | $3.916 \times 10^{-5}$ | $6.718 \times 10^{-5}$ | 65540837,<br>66146213 |

While local ancestry is not likely to be associated with socio-cultural variables, such analyses can still be confounded by population stratification (Mersha 2015). In our study, we opted to include all self-reported Hispanic/Latino individuals in our admixture mapping analysis and adjusted for potential confounding by including global ancestries as covariates in our admixture mapping model. Nevertheless, use of genetically inferred ancestry is pervasive and often heralded as a convenient, albeit arbitrary way of identifying racial/ethnic strata to better control for confounding by population structure (Fang et al., 2019) To evaluate whether our use of self-reported data may lead to starkly different results from using genetically inferred ancestry, we employed the Harmonizing Ancestry Race Ethnicity algorithm (HARE; Fang et al., 2019) to identify individuals whose self-reported race ethnicity labelled may not be consistent with genetically inferred ancestry compared to a reference. Consistent with the goals of our study and best practices in using population labels in human genetic studies^30^, HARE does not attempt to re-assign individuals to strata; rather, individuals with discordant self-report and genetically inferred data are indicated and thus can be excluded from analysis.

Using all available self-reported race/ethnicity labels in the multi-ethnic CCRLP + Kaiser cohort, we observed very low rates of discordance between HARE-predicted labels and the self-reported labels (percent discordance = 0.47% overall, and 0.68%, 3.37%, 0.18%, and 0.54% for AFR, EAS, EUR, and LAT study participants, respectively. **Supplementary Table 14**), suggesting that in general there is not enough genetic evidence to exclude participants from analysis (nor do we advocate for removing individuals from analysis on the basis of genetic make-up alone). In a scenario where individuals might be missing self-reported race/ethnicity labels, HARE can be used to predict the label based on a reference. We simulated such a scenario using 10-fold cross validations, randomly masking in turn 10% of the cohort while using the remaining 90% as the reference for the HARE algorithm to learn the relationship between self-reported labels and genetically inferred ancestry (**Methods**). In this case, we observed the algorithm to be less confident, recalling 61.5% of LAT individuals, compared to 77.5%, 82.6%, and 86.9% of NLW, AFR, and EAS individuals, respectively. Of those not recalled back as Latino, 63.5% were assigned as missing, 35.3% were assigned as Non-Latino White, 0.4% were assigned as Non-Latino East Asian, and 0.8% were assigned as Non-Latino Black (**Supplementary Table 15**). As a sensitivity analysis, we restricted our admixture mapping analysis to the set of Hispanic/Latino individuals inferred by HARE across 10 cross-validation trials, which produced a panel of 1,729 cases and 4,696 controls (**Supplementary Table 16**). Overall, we observed high correlation in odds ratios (Pearson’s correlation coefficient, r = 0. 913) and log *P*-values (r = 0.853), with a modest decrease in significance among variants with *P*-values < 1 x 10^-3^, including the four putative loci (**Supplementary Table 12; Supplementary** Figures 5-6), likely reflecting the smaller sample size.

## Discussion

Motivated by the increased risk of ALL in Hispanic/Latino children, we carried out a systematic investigation into the impact of IA-like ancestry on childhood ALL risk, both at the local and global level. We found that, socio-economic status indicators confound the relationship between global IA-like ancestry and ALL risk, and adjustment for these variables attenuates the association. The lack of global ancestry association, however, did not preclude an association at the local level, as IA-like ancestry was significantly associated with ALL risk across known loci and risk alleles segregated with the IA haplotype more often than expected. On the other hand, we found no interaction between IA-like ancestry and the risk genotype at either the local or global levels, suggesting that there is little evidence the risk alleles exert different effects in different ancestry contexts. Together, these results suggest that, in addition to environmental factors, risk alleles present in higher frequencies in IA populations may contribute to Hispanic/Latino individuals’ excess risk of B-cell ALL.

Further, our in-depth investigation into inferred genetic ancestry also adds to and clarifies previous findings. A previous study found a positive, statistically significant association between global IA-like ancestry and ALL risk^25^. This result may have been affected by selection bias, due to the lower participation of controls than cases in lower SES areas^52^. Associations between global ancestries and disease risks are often reported, but these associations are often attenuated once non-genetic factors are included in the model or when a family-based design is used^53,54^. Indeed, observed associations between ancestry and type-2 diabetes risk in Hispanic/Latinos^26^ and between ancestry and hypertension and lung cancer in African Americans^55,56^ were attenuated once socio-economic covariates were accounted for. In our analysis of the CCLS dataset, we found that the association of ALL risk with global IA-like ancestry was attenuated and no longer statistically significant (though trending in the positive direction) after adjusting for income and education level. We acknowledge that disentangling the association between global genetic ancestry and ALL risk is complex, especially given the observed correlations between SES indicators, ALL risk, and genetic ancestry reported in this study and elsewhere^57,58^. In the CCRLP, however, in which controls were ascertained through statewide birth records^32^ and thus should not be impacted by selection bias, we found no significant association between genetic ancestry and ALL risk.

Our admixture mapping identified associations on chromosomes 2, 7, 10, and 15, but SNPs at the putatively novel loci on chromosomes 2 and 15 did not replicate in additional Hispanic/Latino cohorts. The chromosome 2 locus (2q21.2) is a relative gene desert but overlaps *MGAT5*, which has been reported to be involved in cancer metastasis and tumor suppression in mice^59^. Further, transcriptional regulation of the gene is driven by the Ras-Raf-Ets pathway, which is oncogenic^60–62^. On chromosome 15, the gene closest to the putative risk SNP is *RAB11A*, a member of the RAS oncogene family, and has been previously associated with risk of lung squamous cell carcinoma^63^ and ovarian cancer^64^. Further, this region contains multiple SNP associations with blood cell phenotypes^65,66^, suggesting a possible connection to the etiology of childhood ALL^15^. Adjusting for the lead SNP did not strongly attenuate the chromosome 15 association identified by admixture mapping, suggesting that more complex genomic variation such as structural variants, which may be in low LD with imputed SNPs^67^, could underlie this association and would require further validation. While there may be biological plausibility for these potentially novel loci, the lack of replication could also be due to lack of statistical power, heterogeneity of Hispanic/Latino populations across the United States, or that our initial findings were statistical artefacts.

Within the genetic epidemiology literature, careful consideration has been given to the use of self-reported race/ethnicity or genetically inferred ancestry as strata in performing GWAS. For instance, it is common to define “White British” based on demographic data complemented by principal components of the genotype data when analyzing UK Biobank to remove concerns of stratification^68–70^. However, analysis in a relatively homogenous cohort such as the UKB is not completely free from the effect of stratification^71^, and the notion of defining a homogenous subset of the cohort becomes even more difficult when performing analysis in multi-ancestry cohorts or admixed populations. In those cases, one can pool samples across all ancestries for a “mega-analysis”^72^, or perform stratified analysis based on either (1) self-reported demographic labels^13,32,73^ or (2) some arbitrary heuristics using genetically inferred ancestry^14,18,74,75^. The downside of using genetically inferred ancestry is the prescription of arbitrary thresholds to define ancestry strata. Across studies, the inclusion criteria vary, with individuals of self-reported White, Black and Asian race/ethnicity being defined as study participants with over 75-95% European-like, 50-90% African-like, and 75-90% Asian-like ancestries, respectively^14,18,74–77^.

Previous genetic studies involving Hispanic/Latino participants have defined their strata as individuals whose IA ancestry was greater or equal to 10% *and* greater than the proportion of African ancestry^75,78,79^. Given the heterogeneity of Hispanics/Latinos as a population, this method of categorization may prove untenable. The admixture proportions of Hispanic/Latino individuals vary by country of origin with Caribbean and mainland Hispanics/Latinos exhibiting the highest levels of African-like and IA-like ancestry, respectively^80,81^. Moreover, the estimated admixture proportions can vary depending on the reference samples available, as well as the geographic and temporal definitions of an ancestry component one wishes to estimate. Given these nuances, we have tended to favor relying on self-reported demographic labels as strata when performing multi-ancestry GWAS^32,82^ or when limiting our analyses to Hispanics/Latinos^12^. Further, the results of our HARE analysis showed that the algorithm rarely has enough evidence to exclude an individual from analysis when self-reported labels are present, and imputing missing labels have low recall rate for a heterogenous population like the Latinos. In cases where self-reported labels are missing at high proportions, it may be preferable to conduct a mega-analysis instead of stratified analysis. Indeed, even if overt categorizations impact little of the statistical power of the study design and may provide a bit more robustness to the study, the underlying message conveyed by explicit categorization of people based on estimated genetic ancestry may bring more harm than benefits.

Finally, the results of our study have implications on future investigations of the genetic architecture of ALL and designs of ancestry-based analyses. As reflected in our findings, the elevated ALL risk in Hispanic/Latino children is to some extent driven by the higher frequencies of known ALL risk alleles on IA-like haplotypes, which may underlie the shift towards higher mean polygenic risk scores for ALL in Hispanic/Latino individuals compared to in non-Latino White individuals ^82,83^. Moreover, these consistent directional associations with risk could be indicative of a signature of past adaptation. There is some evidence that ALL risk loci, such as the *IKZF1* locus that we found to be genome-wide significant in admixture mapping, may be under positive selection in Hispanic/Latino populations^12^. Given the overall enrichment of risk alleles on IA haplotypes, the selection may be more oligo-genic or polygenic, though careful scrutiny is needed to rule out other potential confounding^68,84^. Irrespective of whether ALL risk loci are under transient directional selection, the increased prevalence of risk alleles on IA-like haplotypes suggest that continued discovery in larger cohorts with IA-like ancestries and/or utilizing mapping approaches that incorporate signals from both ancestry and allelic associations may be fruitful in discovering more ALL risk loci and in understanding the genetic contributions to risk disparities.

## Data Availability

Genetic data from CCRLP and CCLS were obtained from the California Biobank. Because samples and data are property of the State of California, researchers who would like to request to use the data should establish a protocol with the California Department of Public Health Committee for the Protection of Human Subjects and other relevant agencies to understand if approval to share data are acceptable. The State has provided the following guidance on data sharing, as noted in the following statement: "California has determined that researchers requesting the use of California Biobank biospecimens for their studies will need to seek an exemption from NIH or other granting or funder requirements regarding the uploading of study results into an external bank or repository (including into the NIH dbGaP or other bank or repository). This applies to any uploading of genomic data and/or sharing of these biospecimens or individual data derived from these biospecimens. Such activities have been determined to violate the statutory scheme at California Health and Safety Code Section 124980 (j), 124991 (b), (g), (h) and 103850 (a) and (d), which protect the confidential nature of biospecimens and individual data derived from biospecimens. Investigators may agree to share aggregate data on SNP frequency and their associated p values with other investigators and may upload such frequencies into repositories including the NIH dbGaP repository providing: a) the denominator from which the data is derived includes no fewer than 20,000 individuals; b) no cell count is for <5 individuals; and c) no correlations or linkage probabilities between SNPs are provided." Our data, from CCRLP and CCLS, are each comprised of less than 20,000 subjects and therefore cannot be uploaded to a repository such as dbGaP. Data from Children's Oncology Group and REDIAL were obtained by special request (Yang et al., 2011; Harris et al., 2023). Data from MESA (phs000209.v13.p3) and HCHS/SOL (phs000810.v2.p2.c1, phs000810.v2.p2.c2) were obtained from dbGaP.

## Acknowledgements

We would like to thank the families and patients who participated in each study for ALL, and Soyoung Jeon for helpful conversations contributing to this research. Data used in this study were generated with supports from multiple grants from the National Institutes of Health (R01CA155461, R01CA175737, R01ES009137, P42ES004705, P50ES018172, P42ES0470518, R24ES028524, P20CA262733, U24ES028524, and R01CA262263), the Environmental Protection Agency (RD83615901), the Cancer Prevention and Research Institute of Texas (RP160771, RP210064), and St. Baldrick’s Foundation (consortium grant 522277 with support from the Micaela’s Army Foundation). Analysis performed in this study was supported by F31CA278359 to J.L., R01CA262263 to A.J.D., and R35GM142783 to C.W.K.C. A.J.D. is a Scholar of the Leukemia & Lymphoma Society. The content and conclusions from this work do not reflect the official views of the sponsors. The collection of cancer incidence data used in this study was supported by the California Department of Public Health pursuant to California Health and Safety Code Section 103885; Centers for Disease Control and Prevention’s (CDC) National Program of Cancer Registries, under cooperative agreement 1NU58DP007156; the National Cancer Institute’s Surveillance, Epidemiology and End Results Program under contract HHSN261201800032I awarded to the University of California, San Francisco, contract HHSN261201800015I awarded to the University of Southern California, and contract HHSN261201800009I awarded to the Public Health Institute. The ideas and opinions expressed herein are those of the authors and do not reflect the opinions of the State of California, Department of Public Health, the National Cancer Institute, and the Centers for Disease Control and Prevention or their Contractors and Subcontractors. The biospecimens and/or data used in this study were obtained from the California Biobank Program (CBP request #26 and #1380) Section 6555(b), 17 CCR). The California Department of Public Health is not responsible for the results or conclusions drawn by the authors of this publication. Computation of this work is supported by USC’s Center for Advanced Research Computing (https://www.carc.usc.edu/).

## Data Availability Statement

Genetic data from CCRLP and CCLS were obtained from the California Biobank. Because samples and data are property of the State of California, researchers who would like to request to use the data should establish a protocol with the California Department of Public Health Committee for the Protection of Human Subjects and other relevant agencies to understand if approval to share data are acceptable. The State has provided the following guidance on data sharing, as noted in the following statement: “California has determined that researchers requesting the use of California Biobank biospecimens for their studies will need to seek an exemption from NIH or other granting or funder requirements regarding the uploading of study results into an external bank or repository (including into the NIH dbGaP or other bank or repository). This applies to any uploading of genomic data and/or sharing of these biospecimens or individual data derived from these biospecimens. Such activities have been determined to violate the statutory scheme at California Health and Safety Code Section 124980 (j), 124991 (b), (g), (h) and 103850 (a) and (d), which protect the confidential nature of biospecimens and individual data derived from biospecimens. Investigators may agree to share aggregate data on SNP frequency and their associated p values with other investigators and may upload such frequencies into repositories including the NIH dbGaP repository providing: a) the denominator from which the data is derived includes no fewer than 20,000 individuals; b) no cell count is for <5 individuals; and c) no correlations or linkage probabilities between SNPs are provided.‒ Our data, from CCRLP and CCLS, are each comprised of less than 20,000 subjects and therefore cannot be uploaded to a repository such as dbGaP. Data from Children’s Oncology Group and REDIAL were obtained by special request (Yang et al., 2011; Harris et al., 2023). Data from MESA (phs000209.v13.p3) and HCHS/SOL (phs000810.v2.p2.c1, phs000810.v2.p2.c2) were obtained from dbGaP.

