## Supplementary Figures for "The impact of Indigenous American-like ancestry on risk of acute lymphoblastic leukemia in Hispanic/Latino children"

**Supplementary Figure 1: Ancestry proportions of all cohorts**. The ancestry proportions of CCRLP+Kaiser (**top-left)** CCLS (**top-right**), COG+MESA (**bottom-left**), and ACCESS/REDIAL+HCHS/SOL (**bottom-right**) are shown. Case control status is denoted by "CA" (cases) or "CO" (controls). Proportion of Indigenous-like, European-like, or African-like ancestries are represented by orange, blue, or red coloring, respectively. Global ancestry proportions were calculated by averaging local ancestry assignments, as inferred by RFMIX[36]. The figure was produced using R v.4.3.1[42].

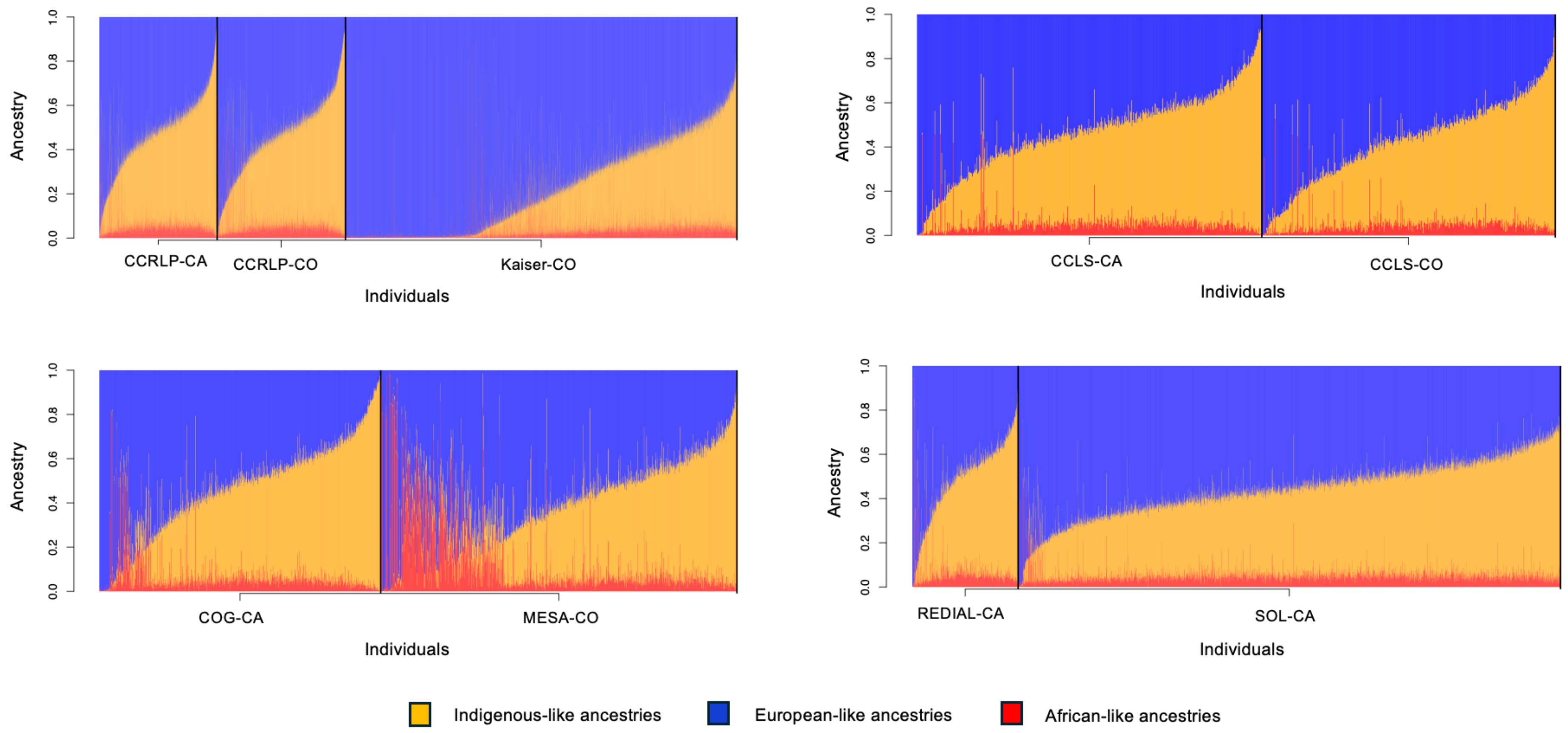

**Supplementary Figure 2: Principal Component Analysis plots of all cohorts**. CCRLP+Kaiser (**top-left**), CCLS (**top-right**), COG+MESA (**bottom-left**), and ACCESS/REDIAL+HCHS/SOL (**bottom-right**) are displayed. Case control status is denoted by “CA” (cases) in red or “CO” (controls) in blue. PCs were inferred using PLINK v2[45] and plotted using R v.4.3.1[42].

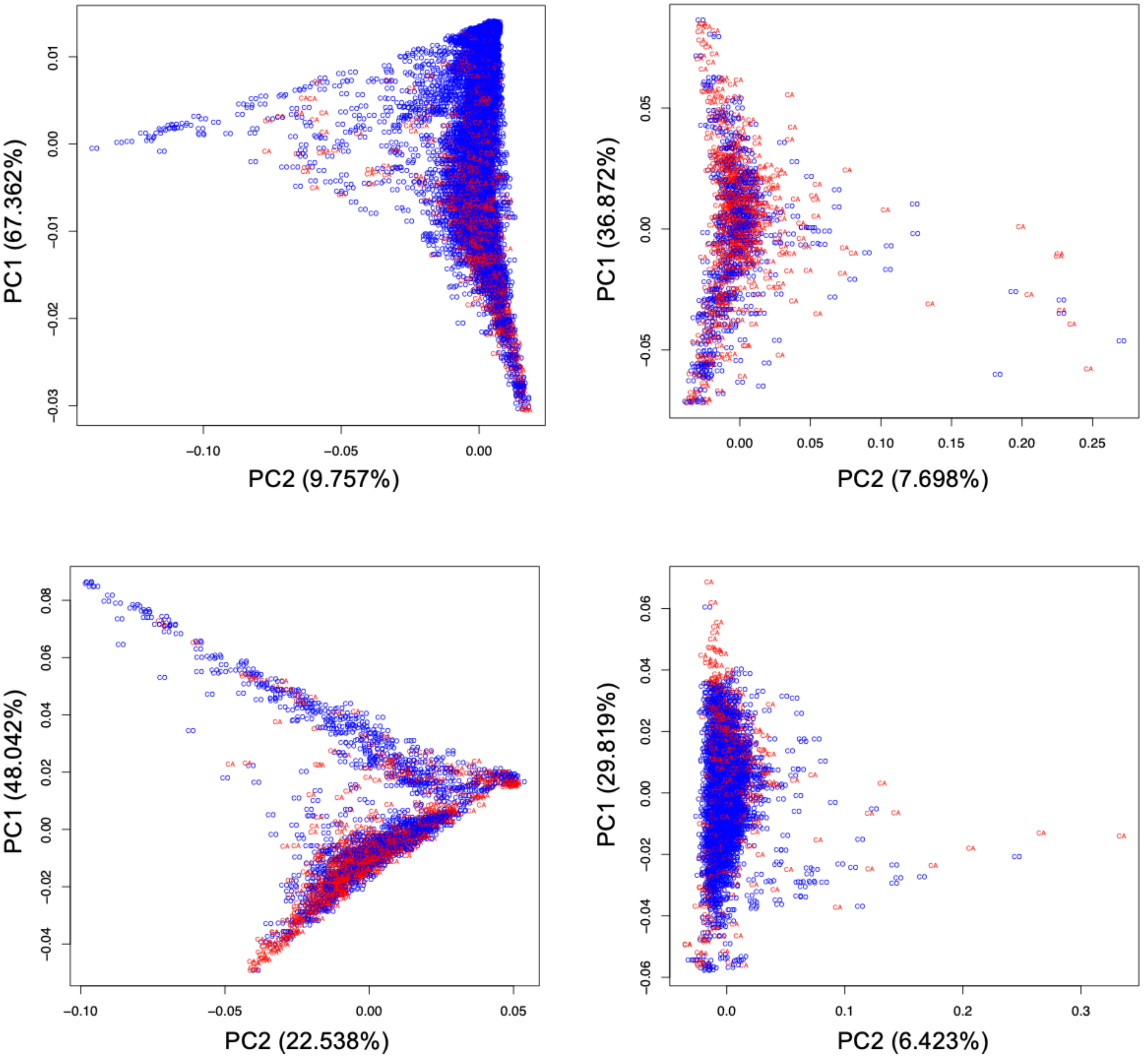

**Supplementary Figure 3: Scatterplot matrix of the first 20 axis from principal component analysis of the multi-ancestry, combined CCRLP and Kaiser data.** Principal component plots of PC1 vs. PC3 and PC2 vs PC3, are enlarged to demonstrate the divergence of East-Asian-like and Indigenous American-like ancestries. Individuals of self-reported Hispanic/Latino, Non-Latino Black, Non-Latino East Asian, and Non-Latino White are represented by yellow, red, green, and blue data points, respectively. Cohort of origin, in the enlarged plots, is denoted as "C" for CCRLP and "K" for Kaiser. The figure was produced using R v.4.3.1[42].

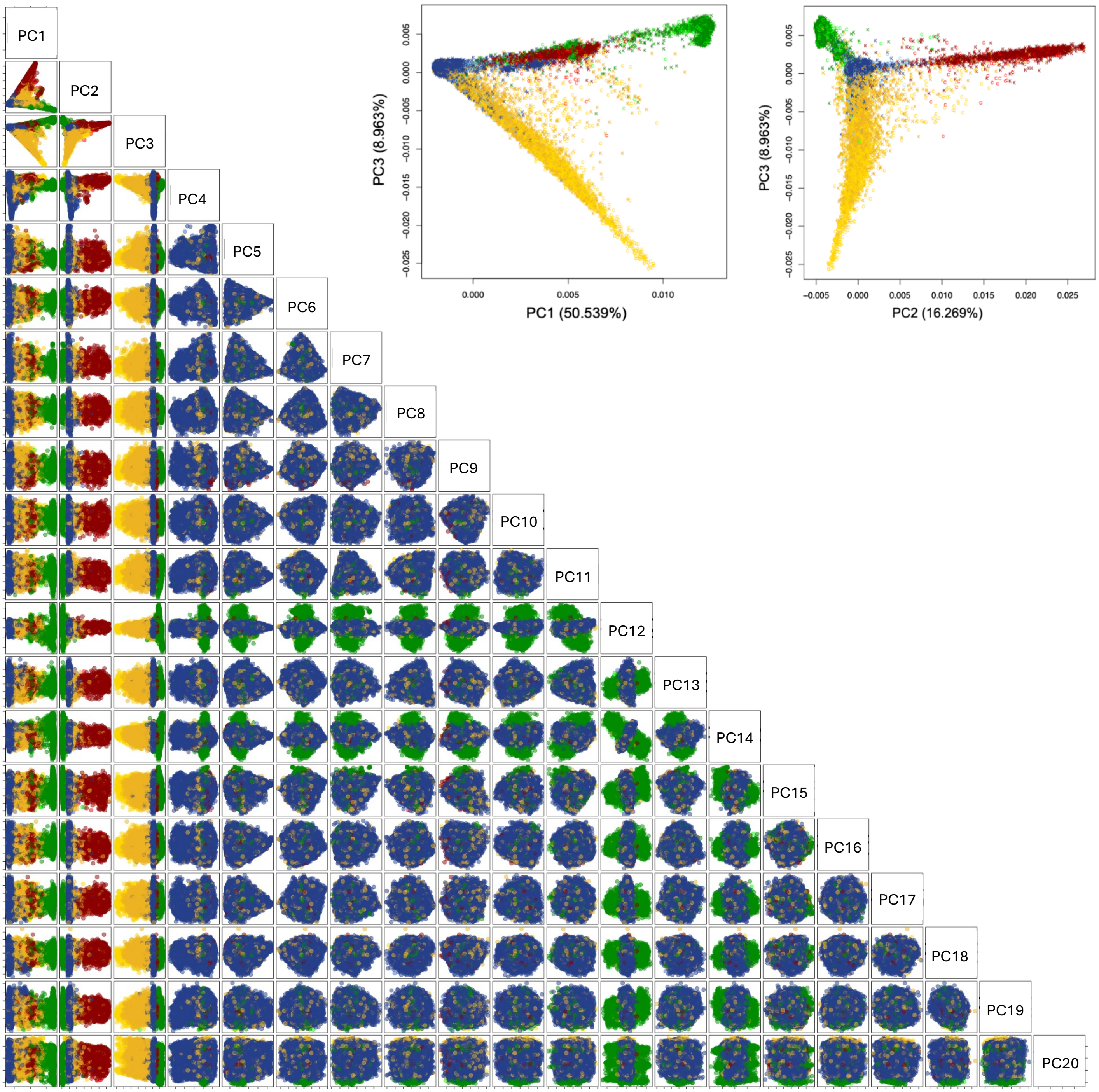

**Supplementary Figure 4**: **Ancestry proportions of those consistently called as Hispanic/Latino across all 10 cross validation trials (LAT) and those who were called as missing or another ancestry group in any of the cross-validation trials (Miscalled).** African-like, Indigenous American-like, and European-like ancestries are depicted in red, orange, and blue, respectively. Global ancestry proportions were calculated by averaging local ancestry, which was inferred using RFMIX[36]. The figure was produced using R v.4.3.1[42].

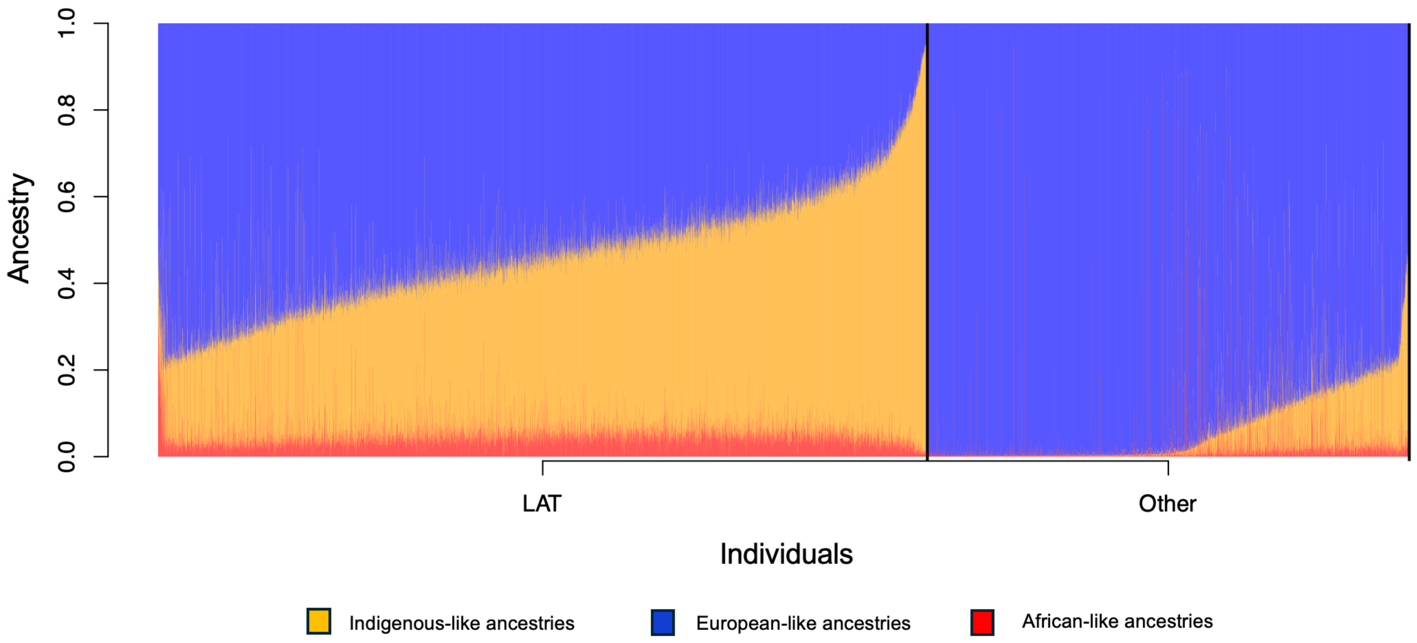

**Supplementary Figure 5: Correlation of -log10(P) values for the admixture mapping discovery analysis using either the full dataset (N = 10450, x-axis) or the reduced dataset with 6,425 individuals who were consistently called as Latinos during 10 cross-validation trials by HARE**. Only SNPs with a p-value < 1 x $10^{-3}$ in either analysis are included. Small jitter to each p-value was introduced prior to log-transformation to improve visualization of SNPs overlapping each other. The plot was produced using R v.4.3.1[42].

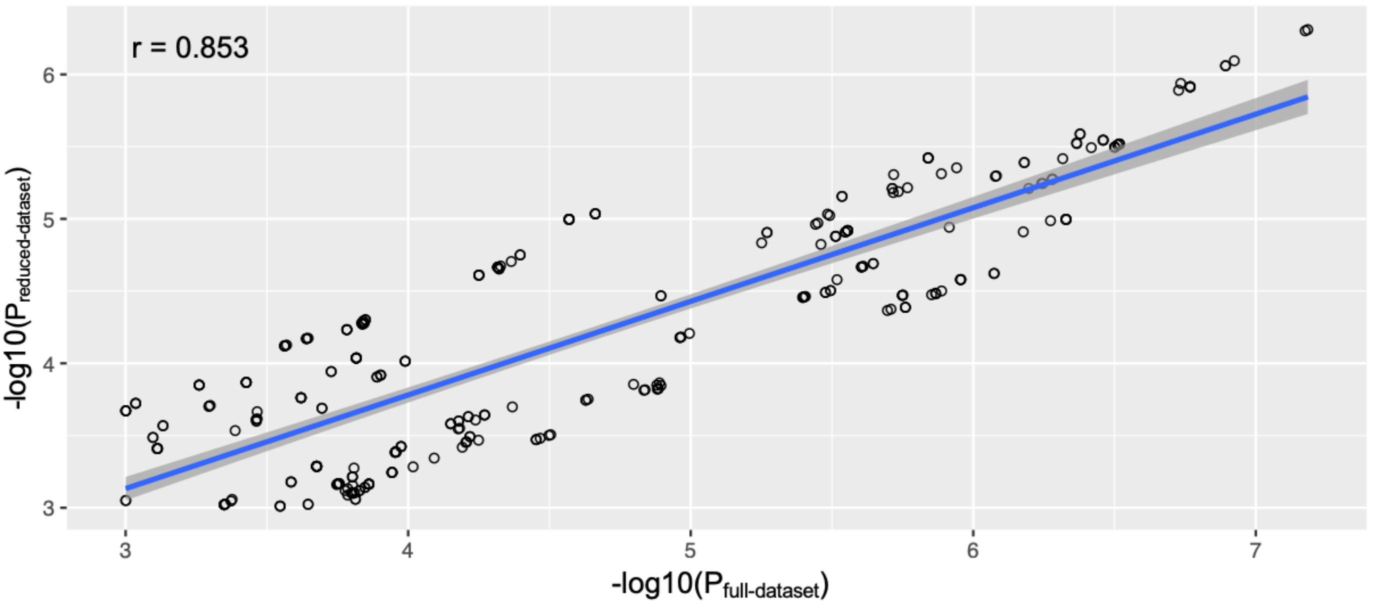

**Supplementary Figure 6**: **Correlation of absolute value of logistic regression coefficients for the admixture mapping discovery analysis either the full dataset (N = 10450, x-axis) or the reduced dataset with 6,425 individuals who were consistently called as Latinos during 10 cross-validation trials by HARE.** Only SNPs with a p-value < 1 x $10^{-3}$ in either analysis are included. Small jitter was introduced to improve visualization of SNPs overlapping each other. The plot was produced using R v.4.3.1[42].

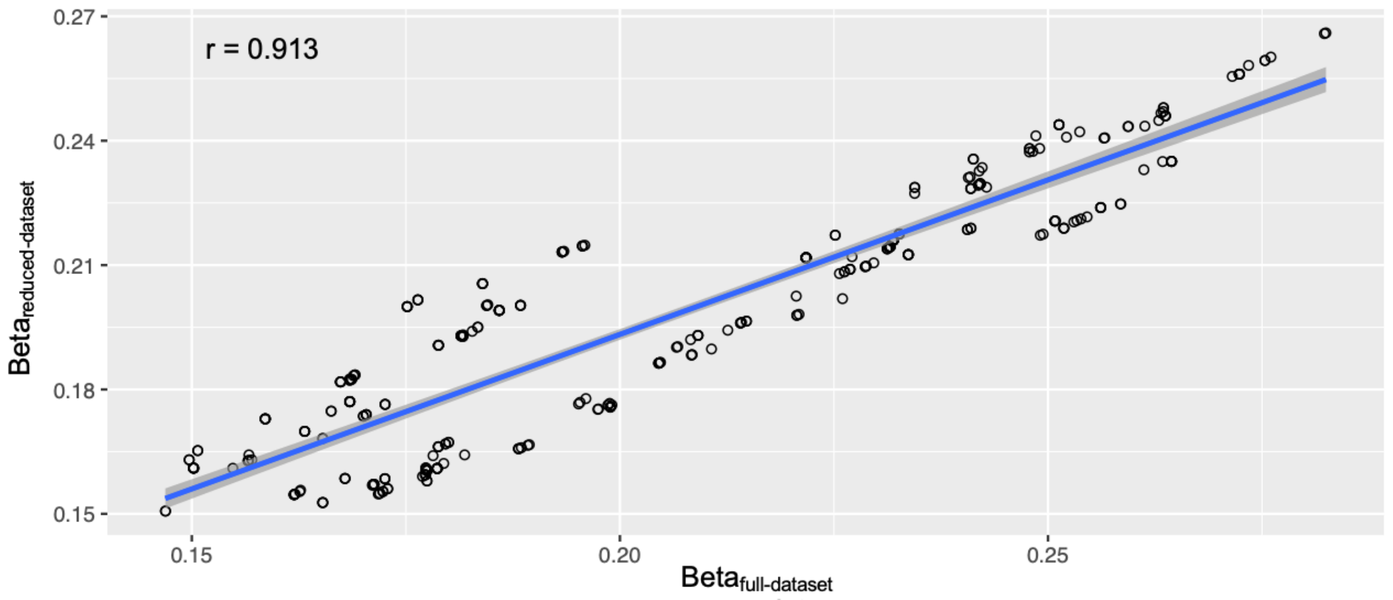
