## Supplementary Tables for "The impact of Indigenous American-like ancestry on risk of acute lymphoblastic leukemia in Hispanic/Latino children"

**Supplementary Table 1**: **The number of remaining individuals and SNPs after Quality Control.** Cases and controls from CCRLP were augmented with controls from Kaiser-GERA. Children's Oncology Group (COG) cases were paired with MESA controls. ACCESS/REDIAL cases were paired with HCHS/SOL controls (from the San Diego recruitment center).

| Study | Total | Cases | Controls | SNPs |
|---|---|---|---|---|
| CCRL + Kaiser-GERA | 10450 | 1930 (CCRLP) | 8520 (2103-CCRLP; 6417-Kaiser) | 536,053 |
| CCLS | 1120 | 605 | 515 | 631,757 |
| COG/ + MESA | 1954 | 836 | 1118 | 523,281 |
| ACCESS/REDIAL+HCHS/SOL | 2295 | 510 | 2285 | 198,974 |

**Supplementary Table 2**: **The association between Indigenous American-like ancestry and ALL case-control status, before and after adjustment for income and parental education, in CCLS and CCRLP**. The Naïve model adjusts for global African ancestry and sex, while the full model adjusts for global African ancestry, sex, and socio-economic status indicators. Odds ratios, the lower (L95) and upper (U95) bound of the 95% confidence intervals, and p-values for each model are shown.

| **Model** | **OR** | **L95** | **U95** | **P-value** |
|---|---|---|---|---|
| $\text{Naïve}_{CCLS}$ | 1.23 | 1.07 | 1.42 | 0.003 |
| $\text{Full}_{CCLS}$ | 1.12 | 0.96 | 1.31 | 0.159 |
| $\text{Naive}_{CCRLP}$ | 1.04 | 0.96 | 1.12 | 0.249 |
| $\text{Full}_{CCRLP}$ | 1.06 | 0.98 | 1.16 | 0.163 |

**Supplementary Table 3**: **The association between household income and ALL case-control status in CCLS and CCRLP.** Odds ratios, the lower (L95) and upper (U95) bound of the 95% confidence intervals, and p-values for each model are shown.

| | **CCLS** | | | | **CCRLP** | | | |
|---|---|---|---|---|---|---|---|---|
| **Income Strata** | **OR** | **L95** | **U95** | **P** | **OR** | **L95** | **U95** | **P** |
| <15,000 | Referent | | | | | | | |
| 15,000-29,999 | 0.76 | 0.53 | 1.11 | 0.153 | 0.75 | 0.48 | 1.18 | 0.213 |
| 30,000-44,999 | 0.57 | 0.38 | 0.85 | 0.005 | 0.77 | 0.49 | 1.19 | 0.24 |
| 45,000-59,999 | 0.54 | 0.35 | 0.83 | 0.005 | 0.79 | 0.50 | 1.25 | 0.321 |
| 60,000-74,999 | 0.30 | 0.17 | 0.52 | $2.130 \times 10^{-5}$ | 0.87 | 0.53 | 1.44 | 0.589 |
| 75,000+ | 0.45 | 0.30 | 0.69 | $2.310 \times 10^{-4}$ | 0.54 | 0.31 | 0.95 | 0.031 |

**Supplementary Table 4**: **The association between parental education on child's ALL case-control status in CCLS and CCLRP**. Odds ratios, the lower (L95) and upper (U95) bound of the 95% confidence intervals, and p-values for each model are shown.

| **Cohort** | **Education Strata** | **OR** | **L95** | **U95** | **P** |
|---|---|---|---|---|---|
| **CCLS** | <High School | Referent | | | |
| | High School | 0.95 | 0.64 | 1.40 | 0.783 |
| | Some College | 0.76 | 0.50 | 1.15 | 0.192 |
| | Bachelors + | 0.73 | 0.46 | 1.15 | 0.173 |
| **CCRLP** | <9$^{th}$ grade | Referent | | | |
| | 9$^{th}$ – 12$^{th}$ grade | 1.21 | 1.00 | 1.48 | 0.055 |
| | Some college | 1.22 | 0.98 | 1.52 | 0.077 |

**Supplementary Table 5**: **The association between estimated global ancestries (African-like, Indigenous American (IA)-like, and European-like) on parental education attainment in CCLS and CCRLP.** The effect size (beta), standard error (SE) and p-values for each model are shown.

| **Cohort** | **Variable** | **Beta** | **SE** | **P** |
|---|---|---|---|---|
| **CCLS** | **Y = African-like** | | | |
| | <High School | Referent | | |
| | High School | 0.003 | 0.005 | 0.560 |
| | Some College | 0.001 | 0.005 | 0.810 |
| | Bachelors + | -0.006 | 0.006 | 0.287 |
| | **Y = IA-like** | | | |
| | <High School | Referent | | |
| | High School | -0.077 | 0.016 | $2.710 \times 10^{-6}$ |
| | Some College | -0.155 | 0.017 | $1.32 \times 10^{-18}$ |
| | Bachelors + | -0.233 | 0.019 | $3.174 \times 10^{-33}$ |
| | **Y = European-like** | | | |
| | <High School | Referent | | |
| | High School | 0.074 | 0.017 | $1.320 \times 10^{-5}$ |
| | Some College | 0.154 | 0.018 | $3.123 \times 10^{-17}$ |
| | Bachelors + | 0.239 | 0.019 | $9.929 \times 10^{-33}$ |
| **CCRLP** | **Y = African-like** | | | |
| | <$9^{th}$ grade | Referent | | |
| | $9^{th}$ – $12^{th}$ grade | 0.004 | 0.003 | 0.090 |
| | Some college | 0.001 | 0.003 | 0.705 |
| | **Y = IA-like** | | | |
| | <$9^{th}$ grade | Referent | | |
| | $9^{th}$ – $12^{th}$ grade | -0.084 | 0.008 | $3.372 \times 10^{-24}$ |
| | Some college | -0.195 | 0.009 | $9.359 \times 10^{-95}$ |
| | **Y = European-like** | | | |
| | <$9^{th}$ grade | Referent | | |
| | $9^{th}$ – $12^{th}$ grade | 0.079 | 0.008 | $8.319 \times 10^{-22}$ |
| | Some college | 0.194 | 0.009 | $2.426 \times 10^{-93}$ |

**Supplementary Table 6**: **The association between estimated global ancestries (African-like, Indigenous American (IA)-like, and European-like) on household income in CCLS and CCRLP.** The effect size (beta), standard error (SE) and p-values for each model are shown.

| | **CCLS** | | | **CCRLP** | | |
|---|---|---|---|---|---|---|
| **Income Strata** | **Beta** | **SE** | **P** | **Beta** | **SE** | **P** |
| **Y = African-like** | | | | | | |
| <15,000 | Referent | | | | | |
| 15,000-29,999 | -0.0003 | 0.005 | 0.946 | -0.003 | 0.006 | 0.659 |
| 30,000-44,999 | -0.001 | 0.005 | 0.791 | -0.007 | 0.006 | 0.223 |
| 45,000-59,999 | 0.007 | 0.005 | 0.214 | -0.007 | 0.006 | 0.276 |
| 60,000-74,999 | 0.003 | 0.007 | 0.678 | -0.012 | 0.007 | 0.078 |
| 75,000> | -0.014 | 0.005 | 0.010 | 0.009 | 0.007 | 0.200 |
| **Y = IA-like** | | | | | | |
| <15,000 | Referent | | | | | |
| 15,000-29,999 | -0.022 | 0.015 | 0.148 | -0.040 | 0.019 | 0.040 |
| 30,000-44,999 | -0.072 | 0.016 | $1.040 \times 10^{-5}$ | -0.074 | 0.019 | $1.130 \times 10^{-4}$ |
| 45,000-59,999 | -0.075 | 0.017 | 1.760 x 10 | -0.109 | 0.020 | $3.260 \times 10^{-8}$ |
| 60,000-74,999 | -0.164 | 0.022 | $1.580 \times 10^{-13}$ | -0.124 | 0.022 | $1.110 \times 10^{-8}$ |
| 75,000> | -0.217 | 0.017 | $9.830 \times 10^{-36}$ | -0.199 | 0.024 | $1.584 \times 10^{-16}$ |
| **Y = European-like** | | | | | | |
| <15,000 | Referent | | | | | |
| 15,000-29,999 | 0.022 | 0.016 | 0.155 | 0.042 | 0.019 | 0.029 |
| 30,000-44,999 | 0.073 | 0.017 | $1.340 \times 10^{-5}$ | 0.082 | 0.0193 | $2.400 \times 10^{-5}$ |
| 45,000-59,999 | 0.068 | 0.018 | $1.520 \times 10^{-4}$ | 0.116 | 0.020 | $5.030 \times 10^{-9}$ |
| 60,000-74,999 | 0.161 | 0.023 | $1.940 \times 10^{-12}$ | 0.136 | 0.022 | $4.490 \times 10^{-10}$ |
| 75,000> | 0.231 | 0.018 | $7.309 \times 10^{-38}$ | 0.190 | 0.024 | $4.390 \times 10^{-15}$ |

**Supplementary Table 7: Table of pruned B-cell ALL risk loci; the full list of known loci was identified via the literature[12].** SNPs were pruned (**bold**) if ancestry dosage was linked between loci (R2 > 0.9) and if they were within 10Mb of each other.

| **Location** | **Gene** | **SNPs** |
|---|---|---|
| 2p16.1 | *BCL11A* | rs2665658 |
| 2q22.3 | *RPL6P5* | rs17481869 |
| 5q31.1 | *C5orf56* | rs886285 |
| 6p21.31 | *BAK1* | rs210143 |
| 6q23 | *MYB/HBS1L* | rs9376090 |
| **7p12.2** | ***IKZF1*** | **rs10272724, rs4917017, rs76880433** |
| 8q24.21 | *CCDC26* | rs4617118 |
| **9p21.3** | ***CDKN2A/2b*** | **rs3731249, rs2811711, rs77728904** |
| 9q21.31 | *TLE1* | rs76925697 |
| 10p14 | *GATA3* | rs3824662 |
| **10p12.2/10p12.31** | ***BMI1/PIP4K2A*** | **rs7088318,rs11591377** |
| 10q21.2 | *ARID5B* | rs7090445 |
| 10q21.3 | *JMJD1C* | rs9415680 |
| 10q21.3 | *TET1* | rs10998283 |
| 10q26.13 | *LHPP* | rs35837782 |
| 12q23.1 | *ELK3* | rs4762284 |
| **14q11.2** | ***CEBPE*** | **rs2239630 ,rs60820638** |
| **17q21.1** | ***IKZF3*** | **rs2290400, rs17607816** |
| 17q21.32 | *IGF2BP1* | rs10853104 |
| 21q22.2 | *ERG* | rs8131436 |

**Supplementary Table 8**: **The odds of each ancestral haplotype harboring the risk allele in CCLS and CCRLP.** Odds ratios, the lower (L95) and upper (U95) bound of the 95% confidence intervals, and p-values through a Chi-squared test for independence for each model are shown.

| | **Haplotype** | **OR** | **L95** | **U9** | **$P_{X}^{2}$** |
|---|---|---|---|---|---|
| **CCLS** | African-like vs Non-African-like | 1.05 | 0.92 | 1.19 | 0.450 |
| | IA-like vs Non-IA-like | 1.33 | 1.26 | 1.41 | $2.614 \times 10^{-23}$ |
| | European-like vs Non-European-like | 0.75 | 0.71 | 0.79 | $5.391 \times 10^{-24}$ |
| **CCRLP** | African-like vs Non-African-like | 1.00 | 0.94 | 1.06 | 0.953 |
| | IA-like vs Non-IA-like | 1.32 | 1.29 | 1.36 | $4.098 \times 10^{-88}$ |
| | European-like vs Non-European-like | 0.76 | 0.74 | 0.78 | $1.170 \times 10^{-85}$ |

**Supplementary Table 9: Attenuation of the admixture mapping signal at known ALL loci when controlling for risk genotype.** Odds ratios, 95% confidence intervals, the admixture mapping and risk genotype adjusted p-value at known ALL loci[11] are shown.

| Gene | Chr | Pos (Hg38) | Location | $P_{AM}$ | $P_{Adj}$ |
|---|---|---|---|---|---|
| BCL11A | 2 | 60599667 | 2p16.1 | 0.138 | 0.138 |
| RPL6P5 | 2 | 145366886 | 2q22.3 | 0.751 | 0.603 |
| C5orf56 | 5 | 132429514 | 5q31.1 | 0.113 | 0.117 |
| BAK1 | 6 | 33579153 | 6p21.31 | 0.208 | 0.373 |
| MYB/HBS1L | 6 | 135090090 | 6q23 | 0.749 | 0.677 |
| IKZF1 | 7 | 50295636 | 7p12.2 | $2.99 \times 10^{-10}$ | 0.161 |
| CCDC26 | 8 | 129143897 | 8q24.21 | 0.084 | 0.703 |
| CDKN2A/2b | 9 | 21970917 | 9p21.3 | 0.115 | 0.102 |
| TLE1 | 9 | 81132456 | 9q21.31 | 0.358 | 0.438 |
| GATA3 | 10 | 8062245 | 10p14 | 0.003 | 0.469 |
| BMI1/PIP4K2A | 10 | 22134373 | 10p12.2/10p12.31 | $5.422 \times 10^{-4}$ | 0.026 |
| ARID5B | 10 | 61961417 | 10q21.2 | $4.924 \times 10^{-9}$ | 0.028 |
| JMJD1C | 10 | 63261130 | 10q21.3 | $5.383 \times 10^{-8}$ | $6.408 \times 10^{-5}$ |
| TET1 | 10 | 68569307 | 10q21.3 | 0.001 | 0.007 |
| LHPP | 10 | 124604740 | 10q26.13 | 0.508 | 0.614 |
| ELK3 | 12 | 96218984 | 12q23.1 | 0.038 | 0.197 |
| CEBPE | 14 | 23120140 | 14q11.2 | 0.022 | 0.727 |
| USP7 | 16 | 8962582 | 16p13.2 | 0.601 | 0.974 |
| IKZF3 | 17 | 39800982 | 17q21.1 | 0.515 | 0.161 |
| IGF2BP1 | 17 | 49014714 | 17q21.32 | 0.728 | 0.347 |
| ERG | 21 | 38417684 | 21q22.2 | 0.7108 | 0.810 |

**Supplementary Table 10**: **Test for SNPxLocal Ancestry interaction.** The effect sizes, standard error, and p-values for the interaction term for genotype and local Indigenous American-like ancestry, after meta-analysis of CCRLP and CCLS, are shown. Analysis for loci with multiple known causal SNPs also included other SNPs within the locus as covariates; these loci are denoted with *.

| Gene | Chr | Pos (Hg38) | Location | rsID | Effect | SE | P |
|---|---|---|---|---|---|---|---|
| *BCL11A* | 2 | 60599667 | 2p16.1 | rs2665658 | 0.002 | 0.059 | 0.973 |
| *RPL6P5* | 2 | 145366886 | 2q22.3 | rs17481869 | 0.062 | 0.205 | 0.764 |
| *C5orf56* | 5 | 132429514 | 5q31.1 | rs886285 | -0.120 | 0.062 | 0.053 |
| *BAK1* | 6 | 33579153 | 6p21.31 | rs210143 | 0.028 | 0.062 | 0.655 |
| *MYB/HBS1L* | 6 | 135090090 | 6q23 | rs9376090 | -0.015 | 0.086 | 0.859 |
| *IKZF1** | 7 | 50295636 | 7p12.2 | rs4917017 | -0.124 | 0.057 | 0.029 |
| *IKZF1** | 7 | 50384350 | 7p12.2 | rs76880433 | -0.026 | 0.076 | 0.729 |
| *IKZF1** | 7 | 50409515 | 7p12.2 | rs10272724 | -0.049 | 0.067 | 0.465 |
| *CCDC26* | 8 | 129143897 | 8q24.21 | rs4617118 | -0.031 | 0.109 | 0.774 |
| *CDKN2A** | 9 | 21970917 | 9p21.3 | rs3731249 | -0.328 | 0.262 | 0.210 |
| *CDKN2A/B** | 9 | 21993965 | 9p21.3 | rs2811711 | -0.048 | 0.146 | 0.742 |
| *CDKN2B** | 9 | 22057531 | 9p21.3 | rs77728904 | 0.069 | 0.187 | 0.712 |
| *TLE1* | 9 | 81132456 | 9q21.31 | rs76925697 | 0.085 | 0.249 | 0.733 |
| *GATA3* | 10 | 8062245 | 10p14 | rs3824662 | 0.064 | 0.057 | 0.256 |
| *BMI1* | 10 | 22134373 | 10p12.31 | rs11591377 | -0.055 | 0.067 | 0.414 |
| *PIP4K2A* | 10 | 22564019 | 10p12.2 | rs7088318 | 0.040 | 0.072 | 0.573 |
| *ARID5B* | 10 | 61961417 | 10q21.2 | rs7090445 | 0.088 | 0.057 | 0.123 |
| *JMJD1C* | 10 | 63261130 | 10q21.3 | rs9415680 | -0.074 | 0.058 | 0.199 |
| *TET1* | 10 | 68569307 | 10q21.3 | rs10998283 | -0.035 | 0.071 | 0.626 |
| *LHPP* | 10 | 124604740 | 10q26.13 | rs35837782 | -0.123 | 0.055 | 0.025 |
| *ELK3* | 12 | 96218984 | 12q23.1 | rs4762284 | 0.036 | 0.057 | 0.528 |
| *CEBPE** | 14 | 23120140 | 14q11.2 | rs2239630 | 0.0400 | 0.056 | 0.479 |
| *CEBPE** | 14 | 23123408 | 14q11.2 | rs60820638 | -0.099 | 0.064 | 0.119 |
| *IKZF3** | 17 | 39800982 | 17q21.1 | rs17607816 | 0.068 | 0.563 | 0.903 |
| *IKZF3** | 17 | 39909987 | 17q21.1 | rs2290400 | 0.020 | 0.055 | 0.719 |
| *IGF2BP1* | 17 | 49014714 | 17q21.32 | rs10853104 | 0.024 | 0.057 | 0.671 |
| *ERG* | 21 | 38417684 | 21q22.2 | rs8131436 | 0.099 | 0.057 | 0.082 |

**Supplementary Table 11**: **Test for SNPxGlobal Ancestry interaction**. The effect sizes, standard error, and p-values for the interaction term for genotype and global Indigenous American-like ancestry, after meta-analysis of CCRLP and CCLS, are shown. Analysis for loci with multiple known causal SNPs also included other SNPs within the locus as covariates; these loci are denoted with *.

| Gene | Chr | Pos | Location | rsID | Effect | StdErr | P |
|---|---|---|---|---|---|---|---|
| *BCL11A* | 2 | 60599667 | 2p16.1 | rs2665658 | -0.084 | 0.272 | 0.759 |
| *RPL6P5* | 2 | 145366886 | 2q22.3 | rs17481869 | 0.800 | 0.678 | 0.238 |
| *C5orf56* | 5 | 132429514 | 5q31.1 | rs886285 | 0.089 | 0.273 | 0.745 |
| *BAK1* | 6 | 33579153 | 6p21.31 | rs210143 | 0.696 | 0.289 | 0.016 |
| *MYB/HBS1L* | 6 | 135090090 | 6q23 | rs9376090 | -0.283 | 0.384 | 0.460 |
| *IKZF1** | 7 | 50295636 | 7p12.2 | rs4917017 | -0.228 | 0.248 | 0.358 |
| *IKZF1** | 7 | 50384350 | 7p12.2 | rs76880433 | -0.122 | 0.298 | 0.683 |
| *IKZF1** | 7 | 50409515 | 7p12.2 | rs10272724 | -0.251 | 0.288 | 0.385 |
| *CCDC26* | 8 | 129143897 | 8q24.21 | rs4617118 | -0.170 | 0.377 | 0.653 |
| *CDKN2A** | 9 | 21970917 | 9p21.3 | rs3731249 | 1.367 | 0.957 | 0.153 |
| *CDKN2A/B** | 9 | 21993965 | 9p21.3 | rs2811711 | -0.090 | 0.522 | 0.863 |
| *CDKN2B** | 9 | 22057531 | 9p21.3 | rs77728904 | 1.134 | 0.650 | 0.081 |
| *TLE1* | 9 | 81132456 | 9q21.31 | rs76925697 | -1.568 | 0.983 | 0.111 |
| *GATA3* | 10 | 8062245 | 10p14 | rs3824662 | -0.264 | 0.254 | 0.298 |
| *BMI1* | 10 | 22134373 | 10p12.31 | rs11591377 | 0.543 | 0.303 | 0.073 |
| *PIP4K2A* | 10 | 22564019 | 10p12.2 | rs7088318 | -0.313 | 0.299 | 0.295 |
| *ARID5B* | 10 | 61961417 | 10q21.2 | rs7090445 | 0.201 | 0.258 | 0.436 |
| *JMJD1C* | 10 | 63261130 | 10q21.3 | rs9415680 | 0.212 | 0.266 | 0.425 |
| *TET1* | 10 | 68569307 | 10q21.3 | rs10998283 | 0.318 | 0.318 | 0.317 |
| *LHPP* | 10 | 124604740 | 10q26.13 | rs35837782 | 0.158 | 0.252 | 0.531 |
| *ELK3* | 12 | 96218984 | 12q23.1 | rs4762284 | 0.206 | 0.258 | 0.424 |
| *CEBPE** | 14 | 23120140 | 14q11.2 | rs2239630 | -0.404 | 0.261 | 0.121 |
| *CEBPE** | 14 | 23123408 | 14q11.2 | rs60820638 | 0.362 | 0.293 | 0.217 |
| *IKZF3** | 17 | 39800982 | 17q21.1 | rs17607816 | 0.326 | 1.682 | 0.847 |
| *IKZF3** | 17 | 39909987 | 17q21.1 | rs2290400 | 0.237 | 0.252 | 0.349 |
| *IGF2BP1* | 17 | 49014714 | 17q21.32 | rs10853104 | 0.385 | 0.253 | 0.128 |
| *ERG* | 21 | 38417684 | 21q22.2 | rs8131436 | 0.562 | 0.284 | 0.047 |

**Supplementary Table 12**: **Replication testing of the putatively associated SNPs responsible for admixture association**. Results were meta-analyzed across CCLS, COG+MESA, and ACCES/REDIAL+HCHS/SOL-SD. Odds ratios, the lower (L95) and upper (U95) bound of the 95% confidence intervals, and p-values are shown.

| Chr | Pos | Ref Allele | Effect Allele | OR | L95 | U95 | SE | P |
|---|---|---|---|---|---|---|---|---|
| 2 | 134116620 | T | C | 0.97 | 0.93 | 1.15 | 0.054 | 0.5129 |
| 7 | 50390453 | C | T | 1.33 | 1.21 | 1.46 | 0.049 | $6.893 \times 10^{-9}$ |
| 10 | 61959980 | T | C | 1.74 | 1.59 | 1.90 | 0.045 | $2.349 \times 10^{-34}$ |
| 15 | 65877726 | G | A | 0.87 | 0.81 | 1.04 | 0.066 | 0.195 |

**Supplementary Table 13**: **Distribution of significantly associated SNPs (with *P* < 0.01) that are in LD with the lead SNP responsible for admixture association**. For each lead SNP on chr2 (rs117345013), 7 (rs6969942), 10 (rs4948492) and 15 (rs12438510), we tabulated the total number of SNPs in LD with them at varying levels of $R^2$ threshold (0.3, 0.5, and 0.8), and the number of those SNPs that showed an association P-value less than 0.01in replication meta-analysis. The lead SNP we tested in replication may be non-significantly associated with ALL due to differences in LD, then we would expect another SNP in LD within the locus should be associated. We found little evidence for chr2 and chr15 to be associated with ALL based on other SNPs in LD.

| | **R2>0.30** | **R2>0.50** | **R2>0.80** |
|---|---|---|---|
| **Chr2** | | | |
| Significant | 0 | 0 | 0 |
| Total | 69 | 39 | 14 |
| **Chr7** | | | |
| Significant | 14 | 13 | 10 |
| Total | 22 | 13 | 10 |
| **Chr10** | | | |
| Significant | 40 | 38 | 15 |
| Total | 40 | 38 | 15 |
| **Chr15** | | | |
| Significant | 1 | 1 | 0 |
| Total | 8 | 6 | 2 |

**Supplementary Table 14: Concordance between self-reported race/ethnicity and HARE strata assignment of CCRLP+Kaiser-GERA study participants.**

| | | SIRE | | | | | |
|---|---|---|---|---|---|---|---|
| | | AFR | EAS | NLW | LAT | <NA> | Total |
| HARE | AFR | 2176 | 0 | 0 | 0 | 0 | 2176 |
| | EAS | 0 | 5155 | 0 | 0 | 0 | 5155 |
| | NLW | 0 | 0 | 58399 | 0 | 0 | 58399 |
| | LAT | 0 | 0 | 0 | 10393 | 0 | 10393 |
| | <NA> | 15 | 180 | 104 | 57 | 0 | 356 |
| | Total | 2191 | 5335 | 58503 | 10450 | 0 | 76479 |

**Supplementary Table 15**: **Distribution of strata assignment of self-reported Hispanic/Latino study participants who were not called back as Hispanic/Latino by HARE, across 10 cross validation trials.**

| Call | Trial 1 | Trial 2 | Trial 3 | Trial 4 | Trial 5 | Trial 6 | Trial 7 | Trial 8 | Trial 9 | Trial 10 | Average |
|---|---|---|---|---|---|---|---|---|---|---|---|
| AFR | 0.009 | 0.010 | 0.010 | 0 | 0.012 | 0.008 | 0.008 | 0.005 | 0.002 | 0.015 | 0.008 |
| EAS | 0.005 | 0.010 | 0.008 | 0.002 | 0 | 0 | 0 | 0.005 | 0.007 | 0.005 | 0.004 |
| EUR | 0.372 | 0.360 | 0.336 | 0.360 | 0.368 | 0.323 | 0.378 | 0.335 | 0.330 | 0.364 | 0.353 |
| NA | 0.614 | 0.620 | 0.646 | 0.637 | 0.620 | 0.669 | 0.615 | 0.655 | 0.659 | 0.616 | 0.635 |

**Supplementary Table 16: P-values of the lead SNPs from admixture mapping using the full dataset (N = 10,450; 1930 cases and 6417 controls) and the reduced dataset (N = 6425; 1729 cases and 4696 controls).** The reduced dataset was restricted to study participants who were called as Hispanic/Latino by HARE consistently across 10 cross-validation trials.

| Chr | Pos | rsID | $P_{Full\text{-}dataset}$ | $P_{Reduced\text{-}dataset}$ |
|---|---|---|---|---|
| 2 | 133605881 | rs2600570 | $4.157 \times 10^{-6}$ | $9.223 \times 10^{-6}$ |
| 7 | 50395237 | rs6952409 | $7.315 \times 10^{-8}$ | $3.030 \times 10^{-6}$ |
| 10 | 62020472 | rs61850777 | $3.210 \times 10^{-5}$ | $6.701 \times 10^{-5}$ |
| 15 | 66111790 | rs12148601 | $3.916 \times 10^{-7}$ | $4.895 \times 10^{-7}$ |
